# Sensor Geometry, Not Signal Processing, Limits Opportunistic Detection of Capillary-Refill-Like Signals by Rule-Based and Language-Model Methods in Archived ICU Waveforms

**DOI:** 10.64898/2026.06.07.26355129

**Authors:** Thomas C. Landry, Youjin Kim

**Author notes:** **Corresponding author** Thomas C. Landry, MD, Internal Medicine, Legacy Salmon Creek Medical Center, 2211 NE 139th Street, Vancouver, WA 98686, USA.

## Abstract

**Background:** Capillary refill time is a resuscitation target in septic shock,^1–4^ but bedside measurement is examiner-dependent. An ICU monitor co-records a photoplethysmogram on the pulse oximeter and intermittent noninvasive blood pressure cuff cycles; if the probe and the cuff share a limb, each cycle is an unplanned vascular occlusion test on the distal microvascular bed. Standard practice places the two on opposite limbs.

**Objective:** To measure how often, in MIMIC-IV-WDB v0.1.0, charted cuff cycles show the photoplethysmographic morphology expected of a same-limb cuff and probe, and to characterize the candidate capillary refill-like signal when that morphology is present.

**Methods:** MIMIC-IV-WDB v0.1.0^5^ was linked to the MIMIC-IV clinical database.^6^ A pre-registered rule-based detector identified candidate occlusion-reperfusion signatures on the 1-Hz perfusion-index envelope around each charted cuff timestamp. The primary endpoint was the proportion of cuff cycles suitable for analysis that were detector-positive at a 15-second reperfusion threshold, with 95% confidence intervals estimated by resampling patients at a fixed seed. A secondary analysis used a locally hosted multimodal language model (a Gemma-3 derivative on an on-device server) to adjudicate the same signature on perfusion-index plots; no MIMIC-IV-WDB content left the workstation.

**Results:** Of 9,224 charted cuff cycles, 8,909 had a usable pulse-oximeter waveform, and 268 cycles in 15 patients (4.30% of the 6,236 cuff cycles suitable for analysis, 95% CI 2.60 to 6.03) met the primary 15-second threshold. The language model adjudicated the same cycles and called 1,367 of the 8,909 cycles with a usable waveform (15.34%) signature-present, roughly five times the detector’s count. Because no laterality ground truth exists, agreement with a single blinded reader served as the comparator rather than accuracy. The two methods were about equally concordant with the reader: precision was 0.25 (95% CI 0.14 to 0.39) for the detector and 0.24 (95% CI 0.10 to 0.35) for the language model, although reweighting to the full population of cycles with a usable waveform lowered the language model to 0.030 (95% CI 0.009 to 0.053). These estimates are reference-limited: a blinded re-read of a 150-card subsample showed only moderate intra-rater reliability (Cohen *κ* 0.46 to 0.59) with systematic undercalling on the first pass, and rescoring against the corrected re-read roughly doubled precision for both methods.

**Conclusions:** Opportunistic extraction of capillary refill-like signals from archived ICU pulse oximetry is limited in two distinct ways. First, sensor geometry limits how often the signal is recordable: cuff cycles rarely show the morphology expected of a same-limb cuff and probe pair, consistent with opposite-limb placement, so the bottleneck is geometry rather than signal processing. Second, the modest reliability of morphology adjudication limits how well any single flagged cycle can be confirmed: against a blinded reader the detector is a usable screen but a noisy confirmer, the reference is itself only moderately reliable, and the language model is no more concordant despite flagging many more cycles. The minority of cycles in which the morphology appears contain a candidate signal that may merit prospective study under controlled placement with laterality recorded.

## 1. Introduction

Capillary refill time is a bedside indicator of distal microvascular perfusion: external pressure briefly empties a small capillary bed, and the time the bed takes to refill on release reflects the integrity of microcirculation. The test is decades old and still routine in emergency and intensive care settings. Two recent multinational randomized trials have reframed it as a resuscitation target in septic shock. ANDROMEDA-SHOCK compared lactate-guided with capillary refill-guided resuscitation in adults with septic shock and reported a non-significant reduction in 28-day mortality with a clearer reduction in organ dysfunction in the refill arm.^1^ A Bayesian reanalysis estimated a high posterior probability of mortality benefit under clinically reasonable priors.^2^ ANDROMEDA-SHOCK-2 then randomized 1,467 adults with early septic shock across 86 centers in 19 countries to a personalized hemodynamic resuscitation protocol targeting capillary refill versus usual care, reporting superiority on a hierarchical composite of mortality, duration of vital support, and length of stay (win ratio 1.16, 95% confidence interval 1.02 to 1.33).^3^ A systematic review with meta-analysis confirmed that prolonged refill predicts mortality across heterogeneous critical-illness cohorts.^4^ The clinical case for an examiner-independent measurement is therefore stronger than before.

Capillary refill belongs to a broader family of microvascular reactivity tests, in which a transient stimulus produces a measurable recovery whose shape reflects vascular function. The vascular occlusion test inflates a cuff above systolic pressure for several minutes and quantifies the post-release recovery, most often using near-infrared spectroscopy on a forearm muscle, and its kinetics correlate with organ-dysfunction severity and outcome in critical illness. Closely related work has applied the same logic to the photoplethysmographic perfusion index after an ipsilateral cuff inflation in septic shock, showing blunted reactive hyperemia and prolonged time-to-peak that tracked illness severity.^7,8^ Capillary refill is a brief, low-amplitude member of this family in which the stimulus is finger compression rather than cuff inflation, and the recovery is observed by eye rather than by an optical sensor. The perfusion index itself, the ratio of pulsatile to non-pulsatile photoplethysmographic components, is a continuous correlate of peripheral perfusion that has been linked to capillary refill and to organ dysfunction.^9^

ICU bedside monitors continuously record a photoplethysmogram on the pulse oximeter probe and intermittently chart oscillometric noninvasive blood pressure measurements from an automatic cuff. MIMIC-IV-WDB v0.1.0^5^ is a credentialed-access subset of these recordings linked to the MIMIC-IV clinical layer^6^ and offers a natural setting to ask a narrow question. If the pulse oximeter probe and the blood pressure cuff sit on the same limb, every routine cuff inflation becomes an unplanned vascular occlusion test on the distal microvascular bed, and the post-release perfusion-index recovery on the photoplethysmogram should resemble the kinetics that deliberate occlusion tests measure. Each ICU patient receives many cuff cycles per day, so a large archived corpus has the potential to yield thousands of opportunistic occlusion-reperfusion events at no incremental cost.

The geometry has to be right. In routine monitoring the pulse oximeter probe and the noninvasive blood pressure cuff are conventionally placed on different limbs because cuff inflation on the same limb transiently occludes flow to the probe, causes pulse-wave loss, and triggers false low-saturation alarms in roughly one in four measurements.^10–12^

The mechanism extends to cuff-pressure transmission at the probe site itself.^13^ No single guideline document mandates opposite-limb placement; the convention lives in device instructions for use and in textbook nursing and anesthesia practice. The same-limb configuration that would deliver the opportunistic capillary refill signal we want is therefore, by design, the off-label condition rather than the routine one. How often it occurs incidentally in a large archived ICU waveform corpus is an empirical question and one not previously reported.

We measured how many charted noninvasive blood pressure cuff cycles in MIMIC-IV-WDB show the photoplethysmographic morphology expected of an ipsilateral cuff and probe pair, asked whether a usable candidate capillary refill signal appears on the photoplethysmogram when that morphology is present (Figure 1), and reported the prevalence as a feasibility estimate. Cuff laterality is not recorded in the data; every detector-positive event is therefore a morphology-based estimate, and we never refer to it as confirmed laterality. We make no outcome claim, no biomarker validation claim, and no clinical prediction model claim. A companion preprint evaluated this cuff-anchored signal alongside two other candidate photoplethysmography-derived microvascular signals for upstream morphology quality.^14^ The present study is the pre-registered, focused analysis of that signal, covering its prevalence, the performance of a rule-based detector, the language-model adjudication, and the blinded-reader reference.

**Figure 1.**
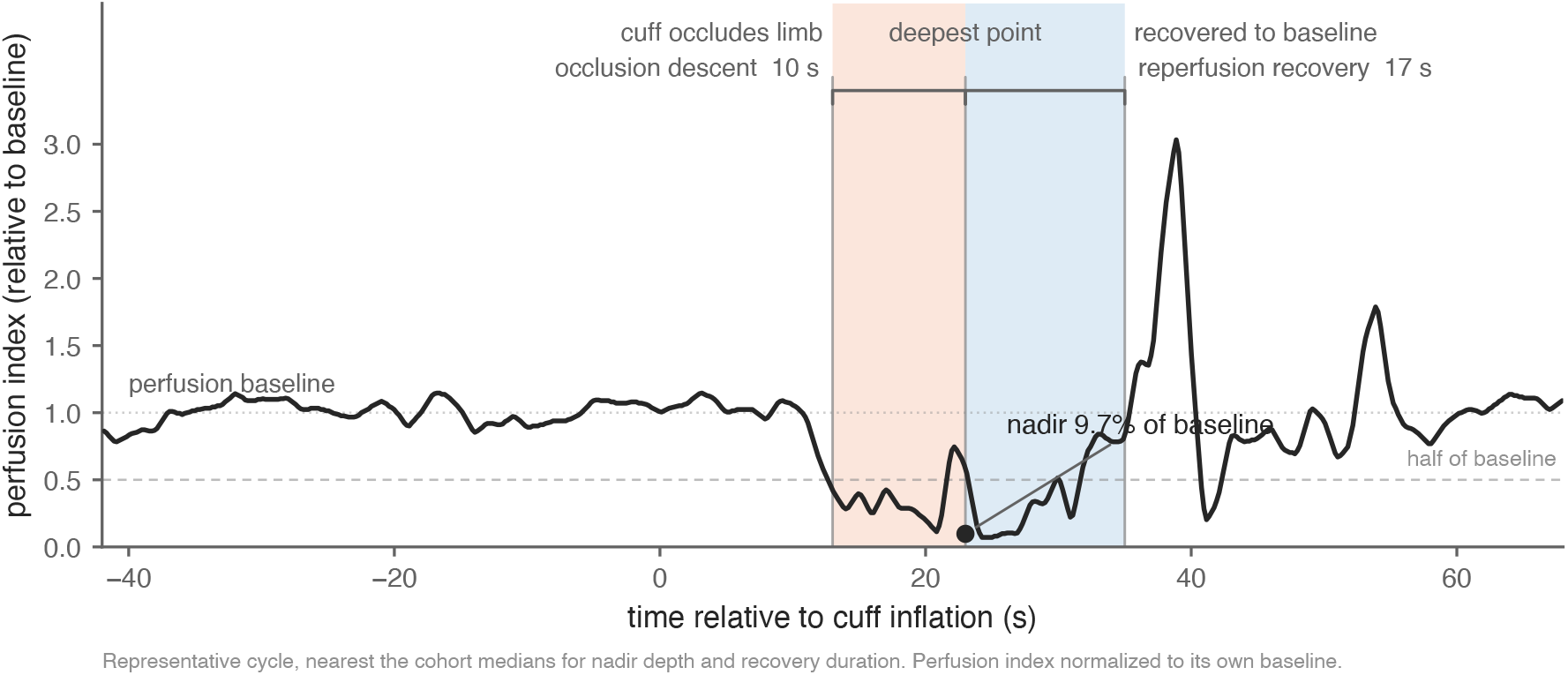
A representative cuff cycle with the perfusion-index morphology expected of same-limb cuff and probe placement. One detector-positive cuff cycle from MIMIC-IV-WDB v0.1.0, selected as the cycle nearest the cohort medians for nadir depth and reperfusion duration among the 268 primary detector-positive cycles. The perfusion index is normalized to its own pre-cuff baseline (baseline = 1.0). The trace shows a stable pre-cuff baseline, a descent to an occlusion nadir near 9.7% of baseline, and a reperfusion climb back toward baseline; shaded spans mark the occlusion descent and the reperfusion recovery, annotated with this cycle’s own measured durations in seconds. The horizontal axis is time relative to the cuff inflation. Cuff laterality is not recorded in the data, so the figure illustrates the candidate morphology and is a morphology-based estimate of same-limb placement, not confirmed laterality.

## 2. Methods

### 2.1. Data sources and linkage

The clinical layer was MIMIC-IV version 3.1^6^ and the waveform layer was MIMIC-IV-WDB v0.1.0, ^5^ both obtained under PhysioNet credentialed access with an executed Data Use Agreement. Downloaded files were SHA-256 verified against the PhysioNet manifest. We linked each waveform record to MIMIC-IV by overlapping the record’s anchored time window with the patient’s ICU-stay window. The linkage yielded 147 unique subjects across 148 ICU stays and 149 waveform records. From this linked set, the analyses reported here used a pilot set of 19 records (19 unique subjects). No MIMIC-IV-WDB or MIMIC-IV content was transmitted off the workstation at any analysis stage.

### 2.2. Cuff event candidates

Each charted noninvasive blood pressure measurement in MIMIC-IV is timestamped with a charted measurement time that PhysioNet describes as a proxy for the moment of measurement; the per-cycle phase (inflation onset, peak pressure, deflation, display update) is not specified.^6^ A cuff cycle was treated as a candidate occlusion-reperfusion event anchored at its charted timestamp. Across the 19 pilot records we identified 9,224 charted cuff cycles, of which 310 had no co-recorded photoplethysmographic window and 5 had a photoplethysmographic window that was more than half missing, leaving 8,909 cuff cycles with a usable pulse-oximeter waveform.

### 2.3. Photoplethysmographic perfusion index and quality control

The primary signal was the photoplethysmographic perfusion index (PI), derived at 1 Hz from the bedside photoplethysmogram channel at its native sampling rate (125 Hz across all records). PI was computed by bandpassing the photoplethysmogram for the pulsatile component, computing its root-mean-square envelope in a rolling window, dividing by a low-passed non-pulsatile envelope, and resampling to 1 Hz. A 5-second rolling median was applied to PI before threshold logic.

A pre-cuff quality-control window spanning 60 to 120 seconds before the charted timestamp required the PI to be plausible (smoothed values between 1% and 100%) and stable (relative standard deviation ≤ 0.30). Of the 8,909 cuff cycles with a usable pulse-oximeter waveform, 6,236 across 18 patients also had a stable pre-cuff baseline and passed this window. We refer to these 6,236 cycles, which had both a usable pulse-oximeter waveform and a stable pre-cuff baseline, as cuff cycles suitable for analysis; they are the primary denominator.

### 2.4. Cuff-occlusion detector: pre-registered parameters

Detector parameters were locked in a pre-registration document^1^ before the yields reported here were computed. The detector operates on the 5-second rolling median of PI around each cuff cycle and identifies four phases of the oscillometric morphology expected when the cuff and probe share a limb: a stable pre-cuff baseline, a deep occlusion in which the smoothed PI runs below 50% of baseline and reaches a nadir below 20% of baseline, a reperfusion envelope from nadir to the time at which PI returns to and stays at or above 85% of baseline for at least 2 seconds, and a return to baseline. The baseline is the median PI over the pre-cuff window. The nadir must lie within an asymmetric alignment window of [− 50, + 30] seconds relative to the charted timestamp; the window is asymmetric because the perfusion-index dip typically leads the charted timestamp. When two or more qualifying runs lie inside the window, the run with the deepest nadir is selected and a flag for multi-dip ambiguity is recorded. A cycle is classified as detector-positive at the primary 15-second reperfusion threshold when all four phase conditions are met and the reperfusion envelope is at least 15 seconds long; a 10-second threshold is reported as a sensitivity stratum (Figure 2).

**Figure 2.**
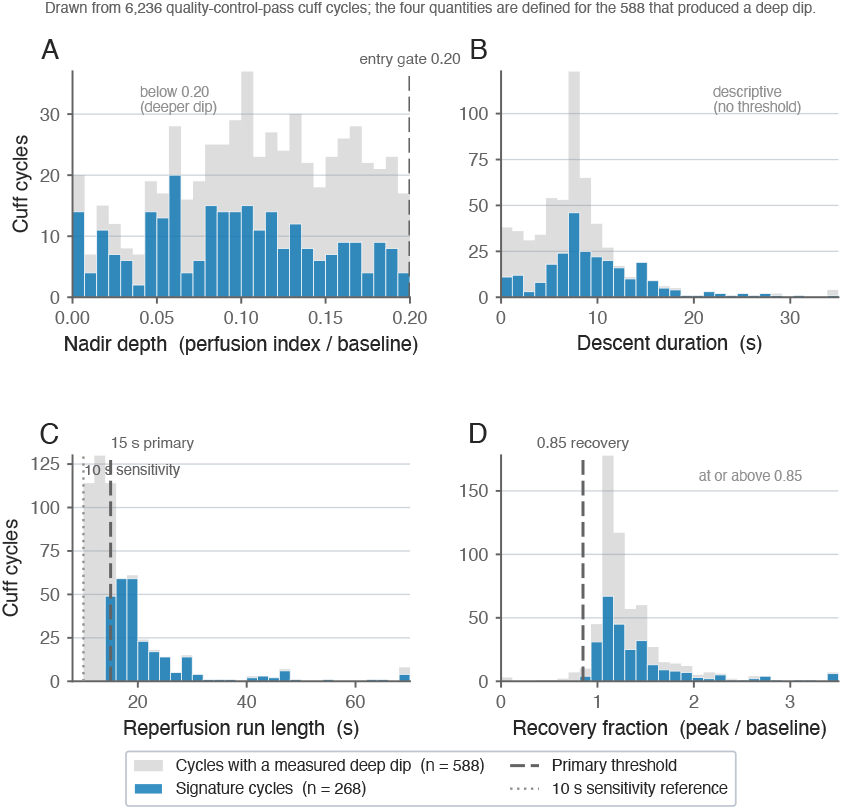
The four measured quantities that define the occlusion-reperfusion signature, with the pre-set decision thresholds. Distributions of the four per-cycle quantities used by the detector, drawn for the 588 cuff cycles that produced a measured deep dip (background) and for the 268 signature cycles (foreground). (A) Nadir depth as a fraction of baseline, with the 0.20 entry threshold. (B) Descent duration, descriptive with no threshold. (C) Reperfusion run length, with the 15-second primary threshold and the 10-second sensitivity reference. (D) Recovery fraction at the end of the analysis window, with the 0.85 recovery threshold. Thresholds are pre-set rule parameters, not values estimated from these data. Counts are morphology-based estimates; cuff laterality is not recorded in the data.

### 2.5. Statistical analysis

The primary estimand was the proportion of cuff cycles suitable for analysis (denominator 6,236) that were detector-positive. Secondary denominators were per-patient (the fraction of records in which at least one detector-positive cycle was observed) and per-all-charted (denominator 9,224). 95% confidence intervals were estimated by resampling patients with replacement, which accounts for multiple cuff cycles per patient, at a fixed random seed (20260426); all cycles belonging to each resampled patient were pooled. A pre-registered subject-clustered split-half analysis split the 19 subjects evenly (9 calibration, 10 held-out) at the global seed, derived a data-driven alignment-window calibration on the qualifying-offset distribution in the calibration half, and applied the pre-registered window plus two data-driven windows (*R*-95 and *R*-90) to the held-out half to estimate sensitivity of the primary yield to the alignment-window choice.

### 2.6. Recovery feature definitions

For each detector-positive cycle the following features were recorded on the normalized PI envelope: depth of nadir (proportion of baseline at the lowest point), reperfusion envelope duration (seconds from nadir to first return to and sustained 85% of baseline), time to 90% recovery, area below baseline during the reperfusion envelope, and overshoot above baseline in the 30 seconds after reperfusion completion. These features are descriptive characterizations of the candidate signal and are not used as outcome predictors in this paper.

### 2.7. Language-model secondary analysis

A locally hosted multimodal language model (a Gemma-3 derivative fine-tuned for medical content, served on an Apple-silicon OpenAI-compatible runtime) read anchor-free PI plots of each candidate cycle and emitted one of three calls (occlusion signature present, no occlusion signature, or indeterminate), with a self-reported confidence value. The prompts were fixed and content-hashed before any inference, and the model identity, prompt version, and run settings were recorded for each call. No MIMIC-IV-WDB content was transmitted off the workstation; the language model ran entirely on the local server. We report results following the TRIPOD+AI^15^ and TRIPOD-LLM^16^ reporting frameworks and the multimodal-AI extension of CLAIM.^17^

Because cuff laterality is not recorded, no laterality ground truth exists against which either the detector or the language model could be scored for accuracy. We therefore framed the comparison as agreement with a human reference rather than as accuracy, following guidance for diagnostic studies with an imperfect or absent reference standard.^18–20^ A single expert reader adjudicated a gallery of perfusion-index cards using the same three calls, blinded to the detector and language-model outputs. To enrich the gallery for the candidate morphology while still spanning the population, cards were drawn by a stratified scheme: a census of all 268 detector-positive cycles, 200 of 320 detector-rejected near-miss cycles, and 100 of 8,107 detector-negative cycles, for 568 cards covering 97.6% of the 8,909 cycles with a usable waveform (Figure 3). This first, complete read (pass 1) is the pre-specified primary reference for the comparison. For each index test we report precision (the positive predictive value, *P* (reader-present | machine-present)), recall or sensitivity (*P* (machine-present | reader-present)), and specificity, scored against pass 1 on the same images, with 95% confidence intervals estimated by resampling patients at the global seed. Because enrichment changes the prevalence of the target and so changes precision,^21,22^ we also reweighted the gallery estimates to the full population of cycles with a usable waveform by the per-stratum sampling fractions (inverse-probability weighting).

**Figure 3.**
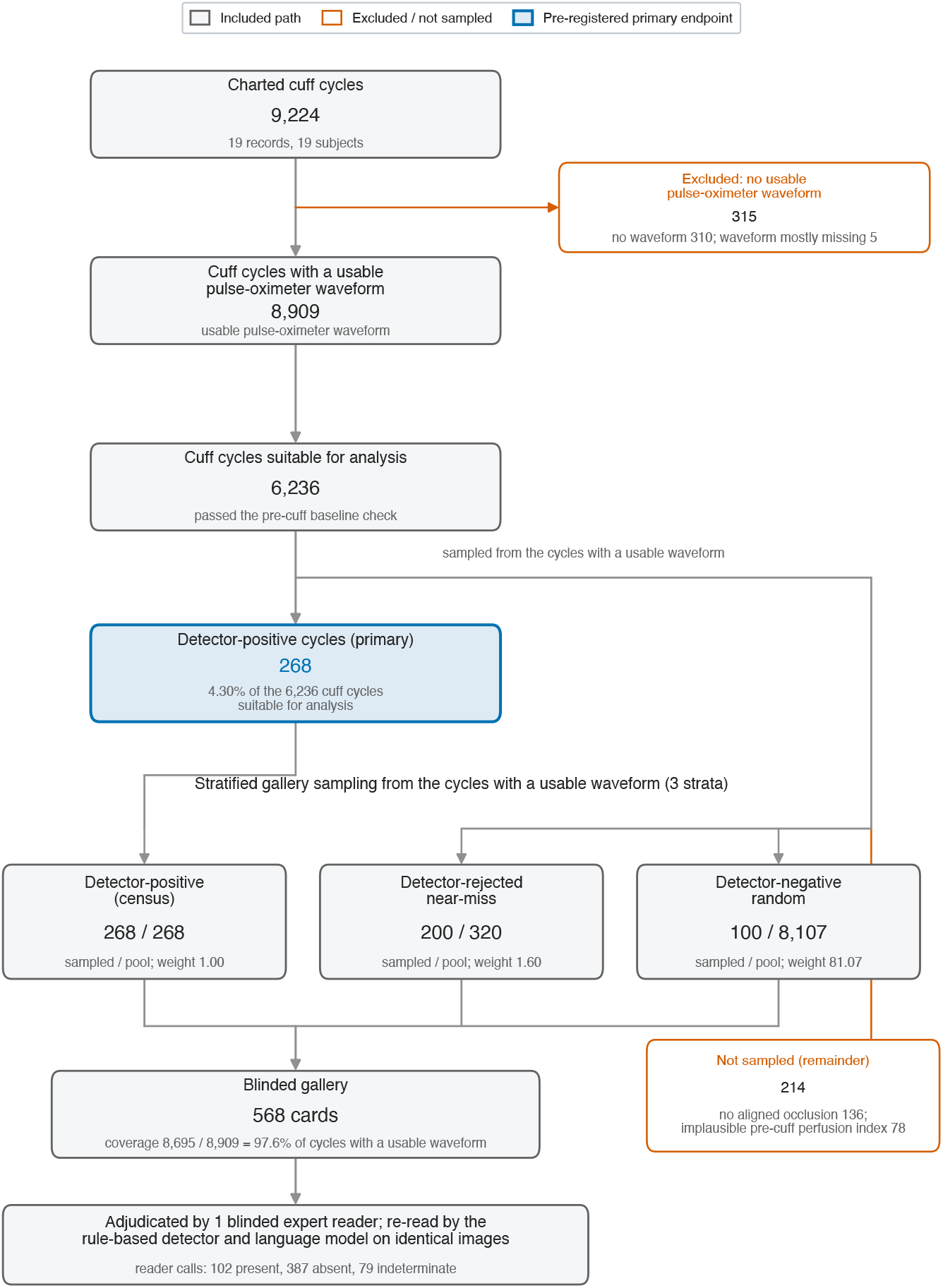
Study flow from charted cuff cycles to the blinded-reader gallery in MIMIC-IV-WDB. Flow of noninvasive blood pressure cuff cycles through screening, detector application, and stratified gallery sampling in MIMIC-IV-WDB v0.1.0. Of 9,224 charted cuff cycles, 315 were excluded for no usable co-recorded photoplethysmographic window (310 with no co-recorded window and 5 with a window more than half missing), leaving 8,909 cuff cycles with a usable pulse-oximeter waveform; 6,236 cycles also had a stable pre-cuff baseline and so are suitable for analysis, and 268 cycles met the primary detector definition at the 15-second reperfusion threshold (4.30% of the 6,236 cuff cycles suitable for analysis, 95% confidence interval 2.60 to 6.03). A 10-second sensitivity threshold yielded 588 cycles. The lower section shows the stratified sampling that built the 568-card adjudication gallery: a census of all 268 detector-positive cycles, 200 of 320 available detector-rejected near-miss cycles, and 100 of 8,107 detector-negative cycles, covering 97.6% of the 8,909 cycles with a usable waveform. The 568 cards were adjudicated by one blinded expert reader, who called 102 cards occlusion-signature-present, 387 no-occlusion-signature, and 79 indeterminate. Counts are morphology-based estimates; the detector cannot confirm cuff laterality from the recorded signal alone.

Because the reader is a single rater, we measured how consistently that reader re-applies the morphology call.^23^ The same reader re-read a stratified 150-card subsample of the gallery, blinded both to the machine calls and to the first-pass calls, and we compared the two passes by percent agreement and Cohen *κ*^24^ with 95% confidence intervals estimated by resampling patients (intra-rater reliability). We then ran a sensitivity analysis on these 150 cards that rescored both index tests against the corrected second-pass calls in place of the first pass, to bound how reference-dependent the precision estimates are. The comparison therefore uses a single-rater reference (pass 1) throughout, with intra-rater reliability reported as the reliability measurement for that reference; inter-rater agreement quantified by an independent second reader is left to future work. A formal latent-class estimate of a notional true call is not identifiable here, because the detector and the language model read the same plotted morphology and so are not conditionally independent given the unobserved laterality, which violates the assumptions of such models.^25^

### 2.8. Reproducibility and code availability

The analysis code is released, and all analyses are reproducible from it. Each stage runs from a documented command-line script with explicit input and output paths and a fixed random seed. A demonstration mode runs end-to-end on the public MIMIC-IV demonstration release without credentialed access. An automated test suite of 278 tests is included, and a content-hashed file manifest records the result files. Detailed commands are provided in the repository. The detector and the perfusion-index pipeline are independent of any language model; the language model is used only in the secondary analysis.

### 2.9. Ethics

Analyses used the Medical Information Mart for Intensive Care IV (MIMIC-IV)^6^ and the MIMIC-IV Waveform Database (MIMIC-IV-WDB v0.1.0), ^5^ both distributed by PhysioNet under credentialed access. The Beth Israel Deaconess Medical Center Institutional Review Board approved the creation and sharing of MIMIC-IV with a waiver of informed consent,^6^ and the authors completed the required human-subjects training and signed the PhysioNet Data Use Agreement before accessing the data. Because the data are fully de-identified and access is governed by the existing institutional approval and the Data Use Agreement, no additional local Institutional Review Board review was required for this secondary analysis. Secondary analytic processing using a locally hosted multimodal language model was performed entirely on a workstation under the same Data Use Agreement; no MIMIC-IV-WDB content was transmitted to any cloud-hosted language model service.

## 3. Results

### 3.1. Cohort and linkage

The linkage of MIMIC-IV-WDB v0.1.0 to MIMIC-IV-3.1 identified 147 unique subjects across 148 ICU stays and 149 waveform records. This analysis used 19 records from 19 unique subjects.

### 3.2. Screening funnel and primary prevalence estimate

Figure 4 summarizes the screening funnel. Of 9,224 charted cuff cycles, 310 had no co-recorded photoplethysmographic window and 5 had a photoplethysmographic window that was more than half missing, leaving 8,909 cuff cycles with a usable pulse-oximeter waveform. The pre-cuff quality-control window admitted 6,236 cuff cycles suitable for analysis across 18 patients. Detector application then rejected 5,512 cycles for the absence of a qualifying deep dip in the alignment window, 136 cycles for a deep dip whose nadir was not aligned with the cuff timestamp, and 302 cycles for a qualifying envelope shorter than the primary 15-second threshold. **Two hundred sixty-eight cycles in 15 patients met the primary detector definition (4.30% of the 6**,**236 cuff cycles suitable for analysis, 95% confidence interval 2.60 to 6.03)**. A 10-second sensitivity threshold added 320 cycles to yield 588 cycles in 15 patients (9.43% of the 6,236 cuff cycles suitable for analysis). Using all charted cycles as the denominator, the primary proportion was 268 of 9,224 cycles, or 2.91%; using the per-patient denominator, 15 of 19 records (79%) showed at least one detector-positive cycle.

**Figure 4.**
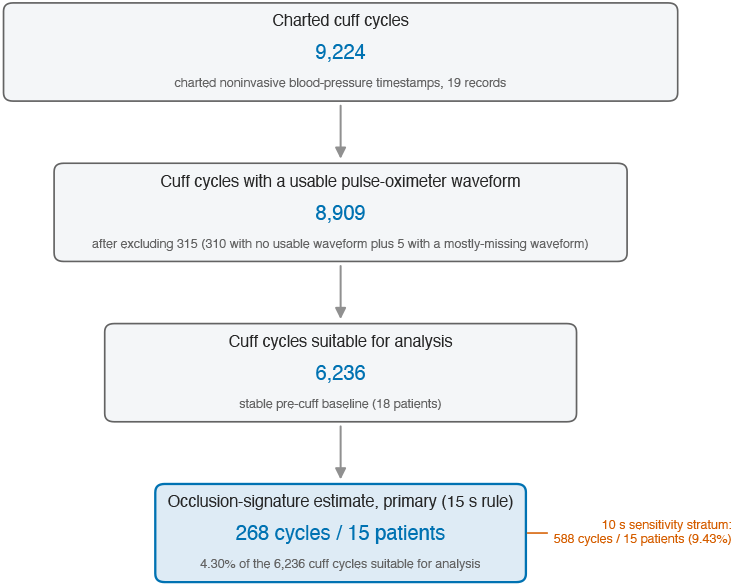
Screening funnel for the cuff-occlusion capillary refill signal in MIMIC-IV-WDB. Counts at each stage of the detector pipeline applied to 9,224 charted noninvasive blood pressure cuff cycles across 19 records (19 unique subjects). Only 315 cycles lacked a usable co-recorded photoplethysmographic window, leaving 8,909 cuff cycles with a usable pulse-oximeter waveform; among these, the largest single exclusion is the absence of a qualifying deep occlusion dip (5,512 cycles). Of the 8,909 cycles with a usable waveform, 6,236 also have a stable pre-cuff baseline and so are suitable for analysis in 18 patients, and 268 cycles in 15 patients show the prespecified occlusion-reperfusion signature on the perfusion-index envelope at the primary 15-second reperfusion threshold (4.30% of the 6,236 cuff cycles suitable for analysis, 95% confidence interval 2.60 to 6.03). A 10-second sensitivity stratum yields 588 cycles in 15 patients. The detector cannot confirm cuff laterality from the recorded signal alone; every detector-positive event is a morphology-based estimate of ipsilateral cuff and probe placement.

### 3.3. Examples of cycles that do not yield a usable signal

Figure 5 displays three representative non-usable cycles, drawn deterministically from the most-represented record for each dominant exclusion stratum. Panel A shows a cycle with no co-recorded photoplethysmographic window. Panel B shows a cycle for which the photoplethysmogram is present but the pre-cuff perfusion-index window is too unstable to set a baseline. Panel C shows a quality-passing cycle on which the perfusion index runs essentially flat through the cuff inflation, the single most common pattern among cycles with a usable waveform and the one consistent with the cuff and the probe being on opposite limbs. Together these three patterns account for the great majority of cycles in this dataset.

**Figure 5.**
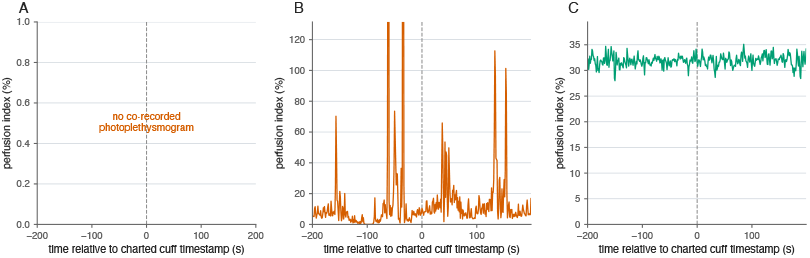
Examples of cuff cycles that do not yield a usable capillary refill signal. Three representative cycles drawn from the dominant exclusion strata. (A) No co-recorded photoplethysmogram available at the charted cuff timestamp. (B) Photoplethysmogram present but the pre-cuff perfusion-index window is unstable, so no baseline can be set. (C) Photoplethysmogram present and pre-cuff window stable, but perfusion index runs essentially flat through the cuff cycle, consistent with the cuff and the probe being on opposite limbs.

### 3.4. A representative candidate cycle

Figure 1 shows a representative detector-positive cycle. The 1-Hz perfusion-index trace and a 5-second rolling median display the four oscillometric phases on the probe-side signal: a stable pre-cuff baseline, a deep occlusion in which the smoothed perfusion index falls well below 20% of baseline, a reperfusion envelope lasting approximately 20 seconds, and recovery to the baseline range. The occlusion dip is closely aligned with the charted cuff timestamp. The depth, duration, and recovery slope are consistent with a post-occlusive reactive hyperemia response on the microvascular bed distal to the cuff. Other detector-positive cycles, including those in a second record that met the 15-second threshold, show the same morphology. Pooling the detector-positive cycles on a common time axis shows a single descent-then-recovery shape with its spread (Figure 6), and the same shape recurs across the patients in whom the signature appears (Figure 7).

**Figure 6.**
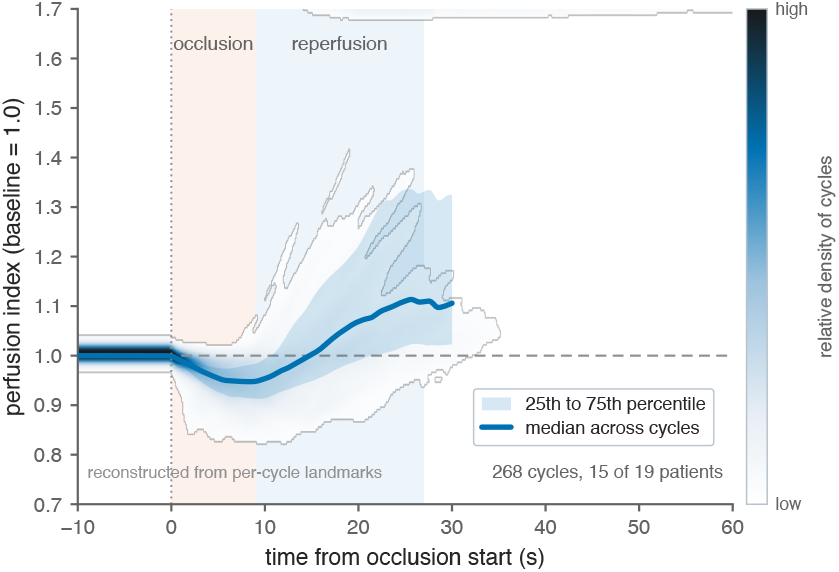
Across-cycle perfusion-index shape of the 268 detector-positive cuff cycles. Each of the 268 primary detector-positive cycles in 15 of 19 records is reconstructed on a common time axis from its measured per-cycle landmarks (baseline, occlusion nadir, and window-end recovery) and normalized to its pre-cuff baseline; the panel shows the density of these reconstructed trajectories with the pointwise median and the 25th to 75th percentile band. The shading marks the occlusion descent and the reperfusion recovery. The trajectories are reconstructed from measured per-cycle landmark distributions, not from resampled waveform samples, and the landmark times are charted on an approximately 1-second grid. *The recovery leg reflects the recovery analysis window (floored at 15 seconds), not a measured time to reperfusion*. Summary curves are computed pointwise over the cycles that still span each instant. All values are morphology-based estimates; cuff laterality is not recorded in the data.

**Figure 7.**
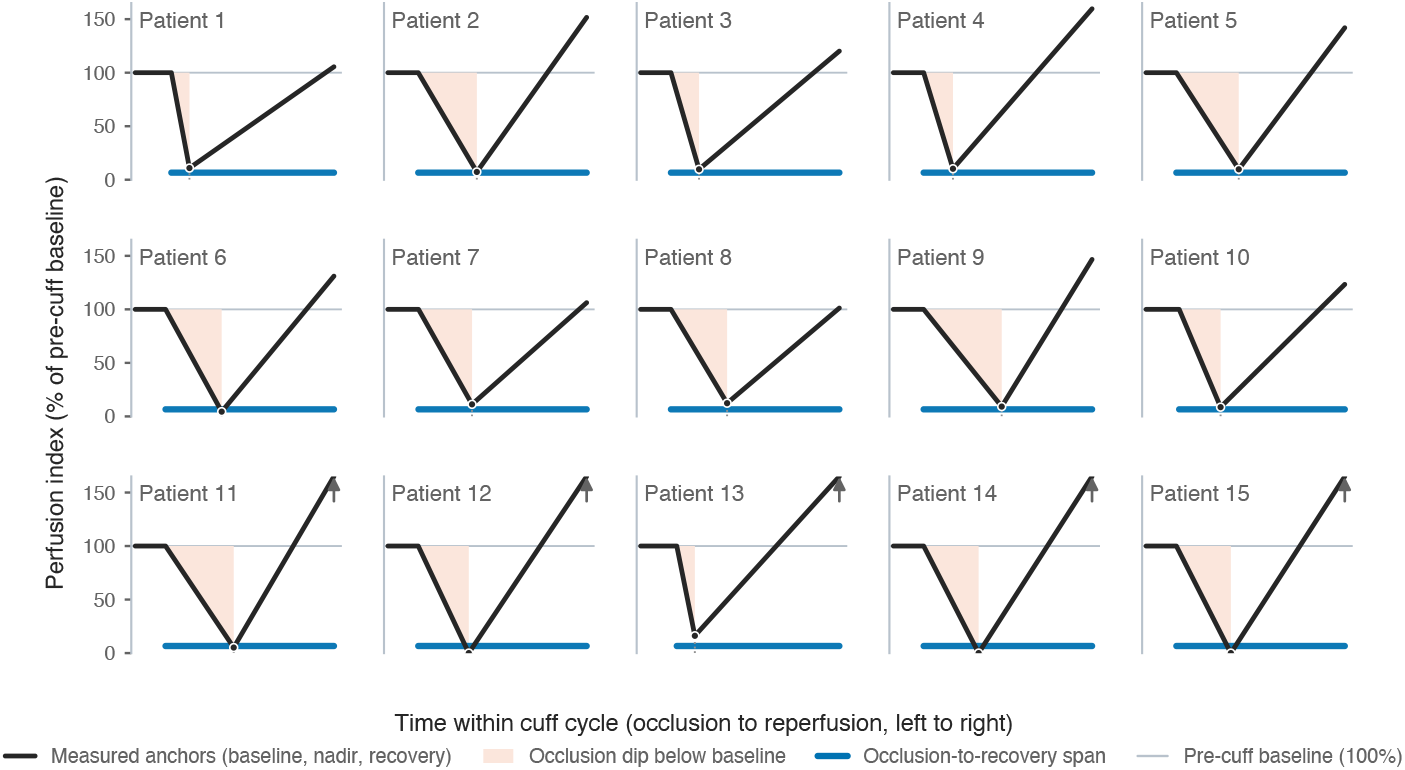
The occlusion-reperfusion shape across the records in which the signature appears. One representative cuff cycle for each of the 15 of 19 records that contained at least one detector-positive cycle; 4 records contained none. For each record the cycle nearest that record’s median nadir depth is shown, drawn as the measured anchor points (pre-cuff baseline, occlusion nadir, and window-end recovery) joined by straight segments; no per-sample waveform is reconstructed. The vertical axis is the perfusion index as a percent of the pre-cuff baseline on a shared scale; an up arrow marks tiles whose recovery overshoots the top of the shared scale. Records are labeled by de-identified ordinal. This is a within-positives view of the records that showed the signature and is not a population prevalence statement; the population estimate is 268 of 6,236 cuff cycles suitable for analysis (4.30%, 95% CI 2.60 to 6.03). All traces are morphology-based estimates; cuff laterality is not recorded in the data.

### 3.5. Sensitivity to detector thresholds and alignment window

A sensitivity analysis on the reperfusion-envelope duration replaced the 15-second primary threshold with a 10-second threshold, retaining the same nadir-depth and alignment constraints; this stratum yielded 588 cycles in the same 15 patients (9.43% of the 6,236 cuff cycles suitable for analysis). The pre-registered subject-clustered split-half analysis split the 19 subjects into a 9-subject calibration half and a 10-subject held-out half at the global seed. The calibration half contained 389 qualifying offsets, with median dip-to-charted-timestamp offset of −9.0 seconds and an interquartile range of [−23.0, +14.0] seconds. The 95th and 90th percentile data-driven alignment windows were [−39.3, +28.0] and [−35.0, +25.6] seconds respectively, both narrower than and contained within the pre-registered [−50, +30] second window. Applied to the held-out half, the pre-registered window yielded 106 cycles in 8 patients (4.41%, 95% confidence interval 1.52 to 9.56%), the data-driven 95% window yielded 4.16% (1.46 to 9.02%), and the data-driven 90% window yielded 3.99% (1.44 to 8.59%). The pre-registered window contained the calibration median and interquartile range, and the pre-registered yield fell inside both data-driven confidence intervals on the held-out half. The primary feasibility estimate is therefore stable to within-cohort calibration of the alignment window.

### 3.6. Secondary comparison: detector, language model, and a blinded reader

The locally hosted multimodal language model adjudicated 8,914 cycles, of which 8,909 had an interpretable waveform (5 could not be parsed, parse rate 99.94%). Of these 8,909 cuff cycles with a usable pulse-oximeter waveform it called 1,367 (15.34%) occlusion-signature-present, 7,542 no-occlusion-signature, and 0 indeterminate. On the same cycles the rule-based detector called 268 positive (3.01% of the 8,909 cycles with a usable waveform), so the language model flagged the candidate morphology roughly five times as often. The full study flow from charted cuff cycles through the stratified gallery to the blinded reader is shown in Figure 3.

A single expert reader adjudicated a stratified 568-card gallery (a census of all 268 detector-positive cycles, 200 of 320 detector-rejected near-miss cycles, and 100 of 8,107 detector-negative cycles), blinded to both machine outputs, and called 102 cards occlusion-signature-present, 387 no-occlusion-signature, and 79 indeterminate. Because no laterality ground truth exists, we scored each method as agreement with this reader rather than as accuracy: precision is the positive predictive value against the reader, and recall is the sensitivity. This first complete read is the pre-specified primary reference. Against it, on the same images, the detector showed a precision of 0.251 (95% CI 0.136 to 0.392), recall of 0.569 (0.458 to 0.810), and specificity of 0.553 (0.461 to 0.660); the language model showed a precision of 0.239 (0.101 to 0.349), recall of 0.539 (0.323 to 0.689), and specificity of 0.548 (0.427 to 0.653) (Figure 8). About one in four positive calls agreed with the reader for each method, so the two were about equally concordant despite the wide gap in how many cycles they flagged. Supplementary Figure S1 shows representative perfusion-index traces for each category of reader-versus-machine call, and Supplementary Figure S2 shows the full three-way pattern of reader, detector, and language-model calls across the 568 cards.

**Figure 8.**
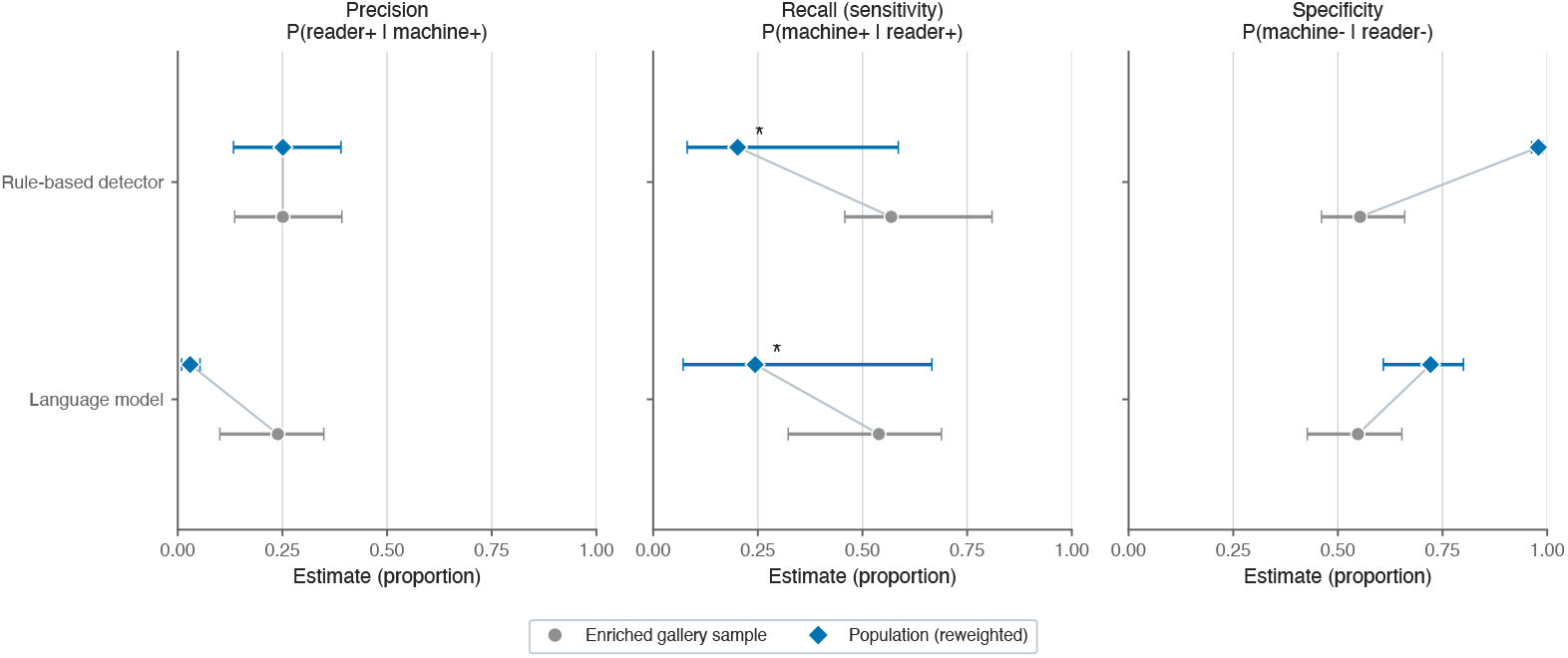
Precision, recall, and specificity of the rule-based detector and the language model against the blinded reader, in the enriched gallery and reweighted to the full population of cycles with a usable waveform. Point estimates with 95% confidence intervals (estimated by resampling patients, seed 20260426) for the two index tests scored against the single blinded expert reader on the same 568 gallery images. Three panels show precision (*P* (reader-present | machine-present)), recall or sensitivity (*P* (machine-present | reader-present)), and specificity (*P* (machine-negative | reader-negative)); the *x*-axis is the estimate as a proportion from 0 to 1. Within each panel the rule-based detector is plotted above the language model. For each index test the enriched-gallery estimate (gray circle) and the population-reweighted estimate (blue diamond) are shown on offset sub-rows joined by a connector; population estimates are reweighted to the full population of cycles with a usable waveform by the per-stratum sampling fractions (inverse-probability weighting, 97.6% coverage). The asterisk in the recall panel marks the two population-recall points, which are driven by only two reader-positive cards in the heavily upweighted detector-negative stratum and have uninformative confidence intervals. All values are morphology-based estimates against a single blinded reader; no laterality ground truth was available.

Because the gallery is enriched for the candidate morphology, precision in the gallery overstates what it would be in the full population, where positives are rarer.^21,22^ Reweighting the gallery estimates to the full population of cycles with a usable waveform by the per-stratum sampling fractions left detector precision unchanged at 0.251 (95% CI 0.133 to 0.390), because the detector-positive stratum is a near-census, but lowered the language model’s population precision to 0.030 (0.009 to 0.053), reflecting its high rate of positive calls on the rarely sampled detector-negative cycles. Precision is therefore prevalence-dependent, and the population figures are the relevant ones for any opportunistic screen applied to an unenriched archive. We report population recall only with caution and do not foreground it, because it rests on just two reader-positive cards in the heavily upweighted detector-negative stratum and has uninformative confidence intervals.

These precision estimates are limited by the reliability of the reference itself. On a blinded 150-card re-read of a stratified subsample, the same reader agreed with the first pass on 66.7% of cards (95% CI 57.4 to 75.4), with a Cohen *κ* of 0.458 on the three-class call and 0.587 for present versus the rest, both moderate.^23,24^The disagreements were directional rather than random. Of the 150 cards, 50 changed call, for a net of 21 additional present calls on the re-read: 23 cards moved to present (8 from absent, 15 from indeterminate), 2 moved from present to indeterminate, and none moved from present to absent. The shift was concentrated in the detector-positive (31 of 71 changed) and near-miss (19 of 53) strata and absent in the detector-negative stratum (0 of 26), where calls were stable. Rescoring both index tests against the corrected second-pass calls in place of the first pass, on the same 150 cards, roughly doubled precision for each method, from 0.250 to 0.500 for the detector and from 0.245 to 0.520 for the language model, while recall was essentially unchanged (0.50 to 0.49 for the detector, 0.43 to 0.51 for the language model) and specificity rose modestly. Precision is thus reference-limited, and the pre-specified first-pass values are conservative. The direction of the change is open to a reader-expectation confound, because the re-read was not blind to the reader’s own prior tendency. Even so, the calls in the detector-negative stratum stayed stable and no card reversed from present to absent, which argues that the re-read corrected first-pass undercalling rather than drifting toward the machines.

Table 1 summarizes the language model’s robustness. Its calls were reproducible across a re-run in a separate session (100 of 100 paired cards in agreement) but sensitive to prompt wording (call concordance 52 to 69% across three minor prompt rewordings on a 100-card subsample) and to how the signal was displayed (38 to 44% concordance against an independently displayed run). The reference here is a single rater, characterized by intra-rater reliability; inter-rater agreement quantified by an independent second reader remains future work. Confidence intervals are wide throughout, consistent with the small number of subjects, and all calls are morphology-based with no laterality ground truth.

**Table 1.**
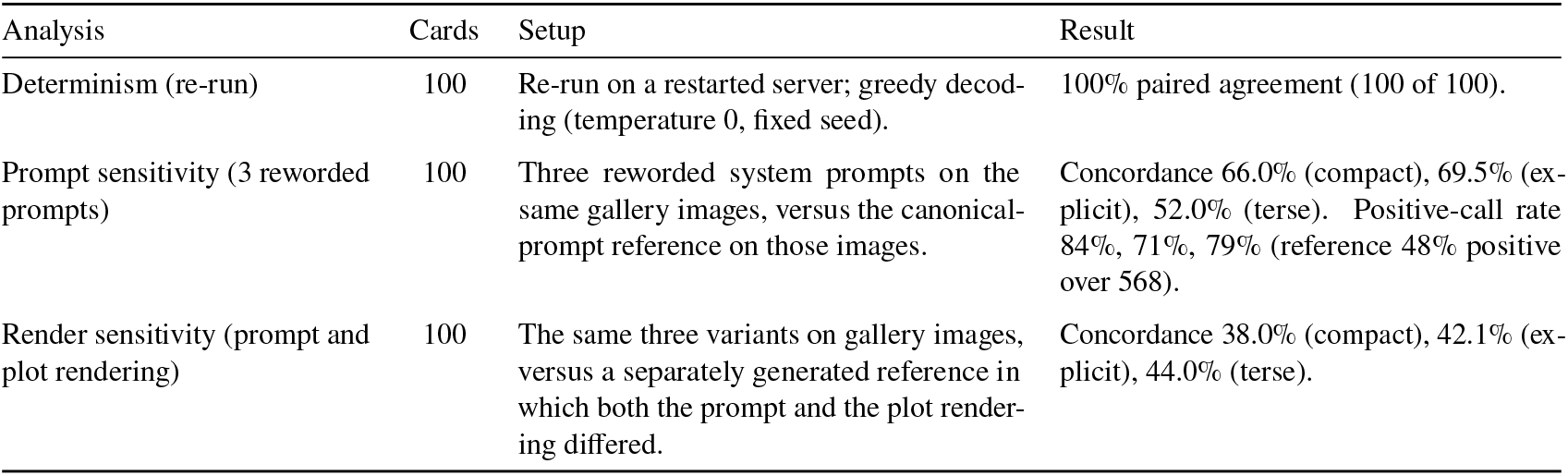
Robustness of the language-model cross-read to re-running, prompt wording, and plot rendering. Three sensitivity analyses of the locally hosted multimodal language model, each on a 100-card subsample of the gallery. The language-model cross-read is an exploratory secondary analysis; all calls are morphology-based estimates.

### 3.7. Recovery feature characteristics of detector-positive cycles

Across the 268 primary detector-positive cycles, the perfusion-index nadir lay at a median of 9.6% of baseline (interquartile range 5.6 to 13.3%), well below the 20% occlusion threshold, and the reperfusion envelope lasted a median of 18.0 seconds (interquartile range 16.0 to 22.0). The perfusion index recovered to a median of 1.24 times baseline by the end of the alignment window (interquartile range 1.09 to 1.51), indicating a modest reactive overshoot above the pre-cuff baseline. The dip-to-charted-timestamp alignment offset had a median of − 11.5 seconds (interquartile range − 24.0 to +8.0), confirming that the perfusion-index nadir typically leads the charted timestamp. We report these features as descriptive characterizations of the candidate signal, not as biomarker estimates.

## 4. Discussion

We screened 9,224 charted noninvasive blood pressure cuff cycles in MIMIC-IV-WDB v0.1.0 and found 268 cycles in 15 patients (4.30% of the 6,236 cuff cycles suitable for analysis) that showed the photoplethysmographic morphology expected of an ipsilateral cuff and probe pair. A 10-second sensitivity threshold roughly doubled the count to 588 cycles, leaving the per-patient denominator unchanged at 15 of 19 records (79%). The primary estimate was stable to alignment-window recalibration on a held-out half of the cohort. When present, the morphology was well-defined: detector-positive cycles showed a stable pre-cuff baseline, a deep perfusion-index nadir, and a 15-to 20-second reperfusion envelope on the same-limb photoplethysmogram. The morphology was absent in the great majority of cycles.

Why so few? The standard-of-care convention is that the pulse oximeter probe and the noninvasive blood pressure cuff occupy opposite limbs, because cuff inflation on the same limb transiently occludes flow to the probe and triggers pulse-wave loss and false low-saturation alarms.^10–12^ The condition we are looking for, ipsilateral cuff and probe placement, is the off-label condition rather than the routine one. Two features of the data are consistent with that interpretation. First, the great majority of cuff cycles did carry a probe-side photoplethysmogram; only 315 of 9,224 cycles (310 with no co-recorded window and 5 with a window more than half missing) were excluded for the lack of one. Among the 8,909 cycles with a usable waveform, the dominant detector rejection was the absence of a qualifying deep occlusion dip in the alignment window (5,512 cycles), followed by an unstable pre-cuff baseline (2,595 cycles). Most cuff cycles therefore show a perfusion index that runs through the inflation without the deep, transient fall that an ipsilateral occlusion would produce, exactly as expected when the cuff and the probe are on opposite limbs (Figure 5C). Second, the 79% per-patient rate at which at least one candidate cycle appears, combined with the 4.30% per-cycle rate, is consistent with a small fraction of qualifying cycles per patient rather than a majority practice in any patient.

When the geometry is right, a candidate signal does appear (Figure 1). The depth, duration, and recovery slope of the perfusion-index envelope on these cycles are consistent with a post-occlusive reactive hyperemia response of the kind that deliberate vascular occlusion tests produce.^7–9^ This shows that the underlying physiology is recordable on routine ICU monitors when the sensors are colocated; it is not proof that the recovered feature is a validated capillary refill measurement. Translating the candidate signal into a clinically useful measurement would require a prospective study with ipsilateral sensor placement, ideally with the cuff laterality recorded in device metadata, and a careful comparison against either the bedside refill test or a microvascular reactivity reference.

A separate question is how confidently any single flagged cycle can be confirmed. Against the pre-specified blinded reader, only about one in four detector-positive cycles agreed with the reader, so the detector is a usable screen but a noisy confirmer of the candidate morphology. That precision roughly doubled, to about one in two, when both methods were rescored against a corrected blinded re-read, and the reference itself was only moderately reliable (intra-rater *κ* 0.46 to 0.59) and systematically undercalled present on first read. Precision here is therefore reference-limited: the bottleneck on confirmation is not only the detector but the reliability of expert morphology adjudication, which is itself a constraint on this kind of opportunistic signal. The multimodal language model was no better, and wrong in a different way.^26^ It flagged roughly five times as many cycles as the detector, yet its concordance with the reader was no higher (gallery precision 0.24 versus 0.25; population precision 0.030 versus 0.251). It tracked the detector’s precision under both the first-pass and the corrected reference, and its calls shifted with prompt wording and with how the signal was displayed while remaining stable on a re-run in a separate session. A latent-class estimate of a notional true call cannot resolve this, because the detector and the language model both read the same plotted morphology and so are not conditionally independent given the unobserved laterality.^18,19,25^

For opportunistic extraction of a capillary refill-like signal from archived ICU waveforms, two separate constraints emerge. The first sets how often the signal is recordable, and it is geometry rather than signal processing: a more capable detector would not change the underlying fact that most cuff cycles in the archive are on the opposite limb from the probe, so the photoplethysmographic occlusion-reperfusion event simply does not occur on the recorded signal. The second sets how well any recorded cycle can be confirmed, and it is the reliability of morphology adjudication: even on the minority of cycles where the event does occur, agreement with a blinded expert reference is modest and that reference is itself only moderately reliable. Future work that aims to derive a capillary refill-like feature from archived ICU waveforms will need either device metadata for cuff laterality, a prospective placement protocol, or a much larger waveform release in which the small fraction of incidentally ipsilateral cycles aggregates to a meaningful sample. It will also need a more reliable adjudication reference, for example multiple independent expert readers or a physiology-anchored standard, before any single flagged cycle can be confirmed. A modestly larger MIMIC-IV-WDB release would, at the per-patient rate observed here, deliver a usable cohort for early prospective comparison work, but only if the count of incidentally ipsilateral cycles per patient is itself stable across releases.

Several limitations bound this report narrowly. The cohort is the 19-record pilot inventory of MIMIC-IV-WDB v0.1.0, an early release described by PhysioNet as 200 records from 198 patients, and the linked-and-extracted analyzable set is small. Cuff laterality is not recorded in MIMIC-IV-WDB; every detector-positive event is a morphology-based estimate, and a counterexample in which a contralateral cuff cycle coincides with a vasomotor dip on the probe-side perfusion index could mimic the signature. The perfusion index is a nonspecific marker of peripheral perfusion that responds to vasomotor tone, temperature, and other factors that vary across patients and over time.^9,27^ The pre-registered alignment window is data-anchored rather than physiology-anchored: MIMIC-IV documents the noninvasive blood pressure charted cuff timestamp as a proxy for the moment of measurement but does not specify the per-cycle phase (inflation onset, peak pressure, deflation end, or display update), so the asymmetric [−50, +30] second window calibrates an empirical offset distribution rather than a known clock relationship.^6^ We treat the asymmetric dip-to-charted-timestamp offset as an empirical finding of this work, supported by mechanistic plausibility from the cuff-deflation phasing literature.^28,29^

The secondary comparison carries further limitations. The reference is a single blinded reader and is therefore an imperfect standard, not a ground truth;^18–20^ its intra-rater reliability was only moderate (*κ* 0.46 to 0.59), and inter-rater agreement quantified by an independent second reader is left to future work. The precision and recall estimates were obtained on an adjudication gallery enriched for the candidate morphology, so the gallery values do not transfer directly to an unenriched archive and we report population-reweighted estimates alongside them.^21,22^ The corrected re-read that raised precision was performed by the same reader and so is open to a reader-expectation confound, although the stability of the detector-negative stratum and the absence of any present-to-absent reversal argue for genuine correction of first-pass undercalling rather than drift. Finally, the cohort is small: the detector inventory spans 19 records and the adjudication gallery spans 16 of the 19 subjects, so all confidence intervals are wide and the secondary comparison is exploratory.

Cuff cycles in MIMIC-IV-WDB v0.1.0 rarely show the photoplethysmographic morphology expected of a same-limb cuff and probe pair. The minority of cycles that do show it carry a candidate signal whose depth and recovery kinetics resemble a post-occlusive reactive hyperemia response, but no validated capillary refill measurement is established here. Opportunistic extraction of capillary refill-like signals from archived ICU pulse oximetry thus runs into two separate limits: sensor geometry limits how often the signal is recordable, and the modest reliability of morphology adjudication limits how well any single flagged cycle can be confirmed. Progress is most likely to come from prospective placement protocols or device metadata that records cuff laterality, paired with a more reliable adjudication reference than a single expert reader.

## Data Availability

The analysis code that reproduces all results is openly available on GitHub (https://github.com/thomas-landry/ipsilateral-cuff-crt-mimic) and archived on Zenodo (https://doi.org/10.5281/zenodo.20588049, version v1.0.0). The underlying datasets, MIMIC-IV (version 3.1) and the MIMIC-IV Waveform Database (MIMIC-IV-WDB v0.1.0), are maintained by the MIT Laboratory for Computational Physiology and distributed by PhysioNet under credentialed access; they are not redistributed here. Investigators can obtain them directly from PhysioNet after completing the required human-subjects training and signing the PhysioNet Data Use Agreement. No patient-level data are contained in the manuscript or the code repository; all derived datasets are regenerable from the released code.

https://doi.org/10.5281/zenodo.20588049

https://physionet.org/content/mimiciv/3.1/

https://physionet.org/content/mimic4wdb/0.1.0/

## Ethics

See Methods, Section 2.9.

## Data and code availability

The analysis code is released, including the locked pre-registration document, the detector parameters, the confidence-interval utility, the split-half alignment-window calibration, the secondary-analysis harness, the fixed prompts, the figure scripts, and the test suite, and is available at: https://github.com/thomas-landry/ipsilateral-cuff-crt-mimic, commit hash 61db1b1868614f324c19ad8ccf4c2f8ea41b719f, archived at Zenodo under version DOI 10.5281/zenodo.20588049 (concept DOI 10.5281/zenodo.20588048, which resolves to the latest version). Each stage runs from a documented command-line script with a fixed random seed, and a demonstration mode runs end-to-end on the public MIMIC-IV demonstration release without credentialed access. An automated test suite of 278 tests is included. The repository pins the software environment and records the result files in a content-hashed file manifest; detailed commands are provided there. MIMIC-IV-3.1^6^ and MIMIC-IV-WDB v0.1.0^5^ are not redistributed; PhysioNet credentialed access and an executed Data Use Agreement are required to reproduce the derived inventories.

## Acknowledgments

MIMIC-IV and MIMIC-IV-WDB are curated by the MIT Laboratory for Computational Physiology and distributed by PhysioNet. The language-model secondary analysis was conducted with a locally hosted multimodal language model; no MIMIC-IV-WDB content left the workstation at any analysis stage.

## Supplementary Material

**Figure S1.**
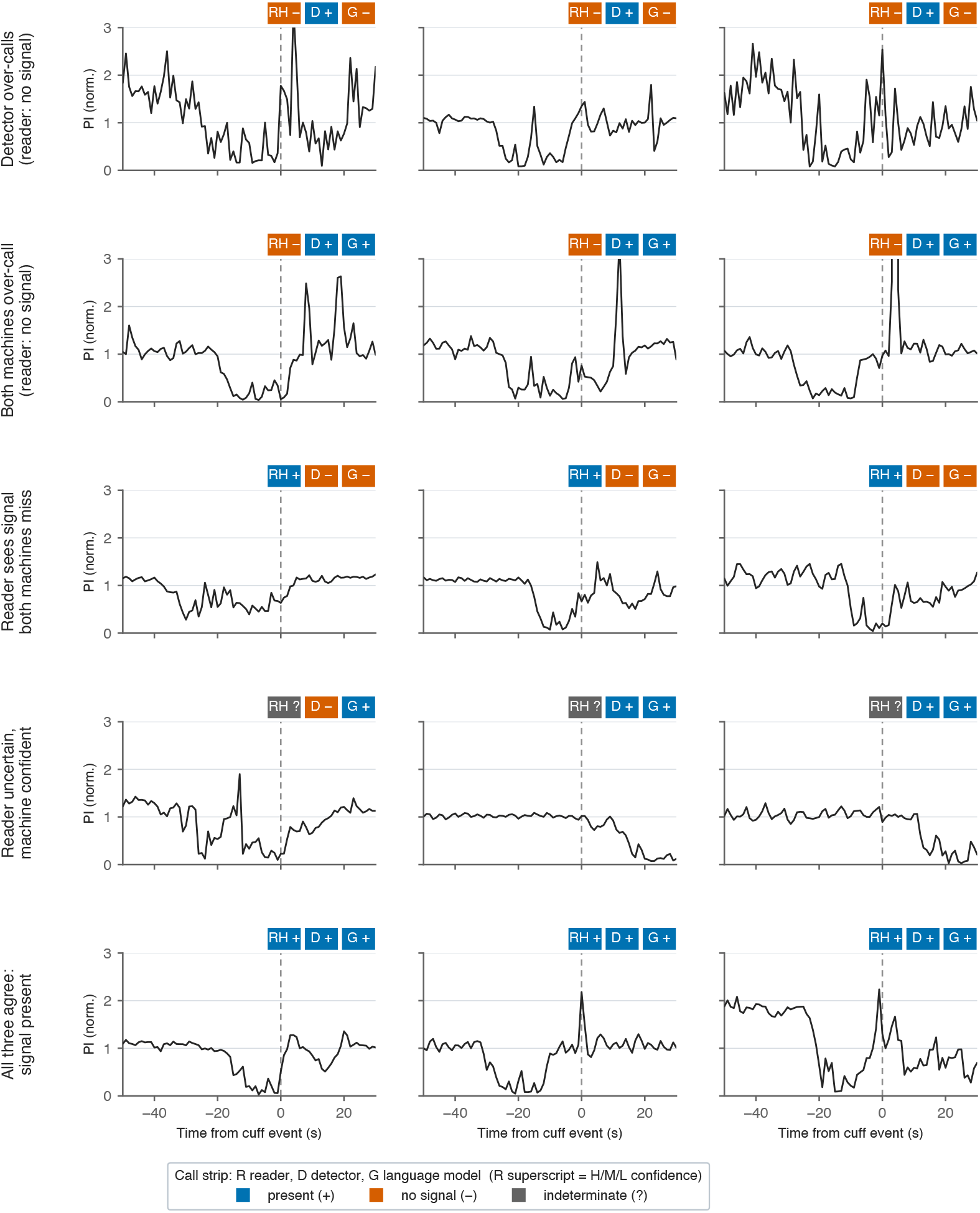
Representative perfusion-index traces by category of reader-versus-machine call. A 5-by-3 grid of perfusion-index traces, one cuff cycle per panel, selected by a prespecified, seeded rule (nearest the per-category centroid in *z*-scored morphology space over reperfusion duration, nadir depth, and alignment offset; seed 20260426) so that each panel is representative rather than hand-picked. Each row is one disagreement category, top to bottom: detector over-calls (reader no-signal, detector present, language model no-signal); both machines over-call (reader no-signal, both machines present); reader sees signal but both machines miss (reader present, both machines absent); reader indeterminate while the language model is confident-present; and all three agree present (a calibration row). Within each panel the *x*-axis is time relative to the charted cuff timestamp in seconds (shared range across all panels, with a dashed reference line at *t* = 0) and the *y*-axis is the median-normalized perfusion index (shared scale across all panels). A three-cell call strip above each panel reports the reader (R), detector (D), and language-model (G) call, colored by call and carrying a redundant glyph (+present, − no signal, ? indeterminate); the reader cell carries a superscript for reader confidence. All 15 cards are drawn from the 568-card gallery. Calls are morphology-based estimates against a single blinded reader.

**Figure S2.**
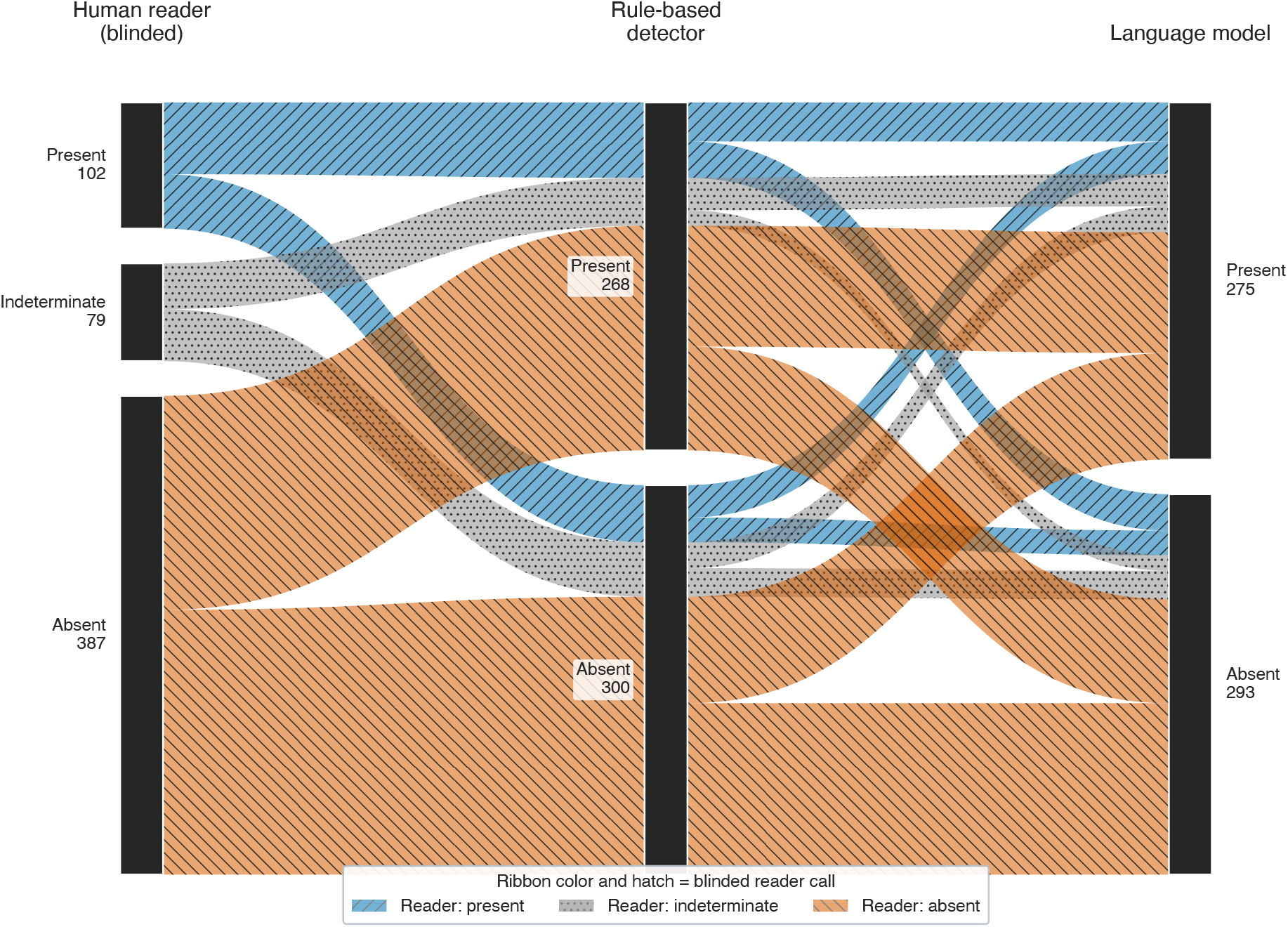
Three-way concordance of reader, detector, and language-model calls across the 568-card gallery. Parallel-categories diagram across three axes, left to right: the blinded human reader (reference), the rule-based detector, and the language model. On each axis the cards are stacked by call; stack heights are card counts. The reader called 102 cards present, 79 indeterminate, and 387 absent; the detector called 268 present and 300 absent; the language model called 275 present and 293 absent (the detector and the language model emit no indeterminate call). Each ribbon is one unique combination of the three calls, its width equal to the number of cards with that combination, colored by the blinded reader call (present, blue; indeterminate, gray; absent, vermillion); the 12 occupied combinations sum to 568 cards. Calls are morphology-based estimates against a single blinded reader; no laterality ground truth was available.

## Footnotes

1 findings/preregistration_detector.md in the project repository; locked 2026-05-22 before any feasibility yield was computed.

